# Posttraumatic stress-related white matter microstructure alterations and chronic pain

**DOI:** 10.1101/2024.07.02.24309869

**Authors:** Yann Quidé, Khaizuran Kamarul, Sylvia M. Gustin

**Author notes:** Corresponding author: Dr Yann Quidé, NeuroRecovery Research Hub, School of Psychology, Biomedical Sciences (Biolink) building, Level 1, UNSW Sydney, NSW, 2052, Australia. These authors contributed equally to this work (Yann Quidé, Khaizuran Kamarul).

## Abstract

**Background:** Posttraumatic stress symptoms (PTSS) are commonly experienced in people with chronic pain. Reduced white matter microstructural integrity in the uncinate fasciculus and the cingulum has been reported similarly in chronic pain and PTSD studies separately. However, the relationship between chronic pain, PTSS and white matter integrity remains unclear.

**Methods.:** Thirty-six subjects with and 20 without chronic pain (controls) underwent diffusion weighted imaging and completed the civilian version of the Posttraumatic stress Disorder CheckList (PCL-C). Average fractional anisotropy (FA) and mean diffusivity (MD) values were extracted from three regions-of-interest (uncinate fasciculus, cingulate and hippocampal portions of the cingulum). A series of multiple linear regressions determined the main effects of Group, PTSS severity (PCL-C total score) and their interactions on each ROI separately.

**Results.:** The group-by-PTSS interaction was significantly associated with variations of uncinate fasciculus FA. Moderation analysis indicated that increasing PTSS severity was significantly associated with reduced uncinate fasciculus FA in the control group, but not in chronic pain group. No other significant association was found for any other ROI FA values, and there was no significant association with MD values.

**Conclusions.:** Consistent with previous studies, increasing PTSS levels were associated with reduced FA of the uncinate fasciculus in controls, but not in people with chronic pain. This suggests that different mechanisms may be at play in chronic pain, including the interplay with other psychopathological problems, such as depression and anxiety. Future larger studies of more homogeneous chronic pain conditions are warranted.

## Introduction

Chronic pain, defined as continuous experience of pain for more than three months or past the expected time of healing (1), is a major public health burden affecting around a third of the worldwide population (2). Chronic pain has severe impacts on both physical (e.g., cardiovascular disease) and mental health (e.g., depression, anxiety), overall affecting the quality of life of people living with chronic pain (3). Trauma exposure, and the development of comorbid posttraumatic stress disorder (PTSD) are strongly related to chronic pain, with around 20% of people with chronic pain experiencing posttraumatic stress symptoms (PTSS), and chronic pain being experienced by 20% to 80% of people diagnosed with PTSD (4, 5). Chronic pain and PTSD are associated with overlapping morphological brain alterations of stress-sensitive brain regions, including the hippocampus, amygdala, and medial prefrontal cortex (mPFC) (6). A recent study has identified other regions sensitive to PTSS in chronic pain that included the middle cingulate cortex (MCC), and the posterior insula (7). These regions are physically connected through common white matter tracts, including the cingulum and uncinate fasciculus (8). However, the impacts of PTSS on these critical white matter tracts in chronic pain remain unclear.

The medial temporal lobe, including the amygdala and hippocampus, is physically connected to the mPFC via two distinct pathways. The cingulum is divided in two portions: the *hippocampal* portion (CGH) connecting the hippocampus to the posterior cingulate cortex (PCC), and the *cingulate* portion of the cingulum (CGC), connecting the PCC to the mPFC via the MCC and the dorsal anterior cingulate cortex (8). On the other hand, the uncinate fasciculus is a shorter, direct tract physically connecting the amygdala in the medial temporal lobe, and the mPFC (8).

Across various chronic pain types (e.g., nociceptive, neuropathic pain) and locations (e.g., orofacial, lower body), people with chronic pain commonly display alterations in white matter fractional anisotropy (FA), a marker of microstructural integrity (9). Compared to control groups (without chronic pain), reduced cingulum and uncinate fasciculus FA has been commonly reported. For example, participants with chronic neuropathic pain following spinal cord injury presented with reduced cingulum and uncinate fasciculus FA (10, 11). People with chronic trigeminal neuralgia, specifically unilateral (right side) orofacial pain, presented with lower cingulum FA than their control group (12). Similarly, lower FA for both the cingulum and the uncinate fasciculus were observed in individuals with chronic temporomandibular disorder (13). However, other studies have reported no differences in white matter microstructure integrity in any of these tracts (14–17), suggesting that chronic pain alone may not be the only factor contributing to these alterations in white matter integrity.

Reduced FA in the same white matter tracts has been reported in PTSD or in association with PTSS (18, 19). In a study with trauma-exposed individuals, PTSS severity was negatively associated with lower uncinate fasciculus FA (one-month post-trauma) and the cingulum (three months post-trauma) (20). Using a whole brain tract-based spatial statistical approach, decreased integrity of the posterior cingulum was observed in a study with African American women with PTSD (21). However, another study also reported no association between white matter integrity changes and PTSD symptoms, in a sample of traumatically injured patients (22). Together, the above studies indicate a large overlap of changes in white matter microstructure on the cingulum and the uncinate fasciculus in both chronic pain and PTSD. However, the impact of PTSS on white matter integrity in people with chronic pain remain unclear.

The present study sets out to characterise the role of comorbid PTSS on white matter microstructure integrity of people with chronic pain. We hypothesised that the relationship between white matter integrity in the cingulum (CGC, CGH) and uncinate fasciculus (FA and mean diffusivity, MD), and variations in PTSS would be different in people with chronic pain compared to controls without chronic pain. We expected that the cumulative effect of chronic pain and PTSS would be evident in lower FA and larger MD values in these tracts for people with chronic pain compared to controls, even those reporting more severe PTSS.

## Methods

### Participants

Participants were 36 people with chronic pain conditions lasting for more than three months (together referred to as the *chronic pain* group), including temporomandibular disorder (n=14), trigeminal neuropathic pain (n=8), burning mouth (n=1), trigeminal neuralgia (n=6), temporomandibular disorder + trigeminal neuropathic pain (n=1), and spinal cord injury neuropathic pain (n=6; complete paraplegia with continuous burning and/or shooting pain in areas of sensorimotor loss), as well as 20 people without chronic pain (controls, HC). Inclusion criteria for all participants were age over 18 years old with no known diagnosis of psychiatric disorder. Exclusion criteria included heart pacemaker, metal implants, intrauterine contraceptive device, insulin pump, infusion devices, hearing-disease, claustrophobia, pregnancy, a history of stroke, multiple sclerosis, or Parkinson’s disease. All participants were volunteers who provided informed consent according to procedures approved by the Human Research Ethics committees of the University of New South Wales (HC15206), the University of Sydney (HREC06287) and Northern Sydney Local Health District (1102-066M).

### Assessments

The civilian version of the PTSD Checklist (PCL-C) (23) is a standardized self-report 17-item questionnaire used to measure the severity and burden of PTSS. Participants indicate how much they have been bothered by a symptom over the past month using a 5-point scale (1 = not at all, 5 = extremely). In this study, no provision of a formal PTSD diagnosis was intended. The total score to the PCL-C (PCL-C total score, ranging from 17 to 85) was used first in analyses, and associations with the severity of specific symptoms were then explored: re-experiencing (cluster B; questions 1-5, ranging from 5 to 25), avoidance (cluster C; questions 6-12, ranging from 7 to 35) and hyperarousal symptoms (cluster D; questions 13-17, ranging from 5 to 25). Severity of depressive symptoms was measured using the Beck Depression Inventory (BDI-I total score; ranging from 0 to 63) (24), and the severity of state anxiety was assessed using the State Anxiety Inventory (SAI total score; ranging from 20 to 80) (25).

Participant’s pain intensity was measured using a visual analogue scale (VAS) in two ways. First, ‘pain diary’ index was used reporting participants experienced levels of pain on a 10-cm horizontal ruler (‘no pain’ = 0; ‘worst pain imaginable’ = 10) three times a day (morning, noon, and evening) for seven days prior to their visit to the scanner. Second, ‘scan pain’ index was used to measure pain intensity during scanning (Table 1).

**Table 1.** Sociodemographic and clinical characteristics of the studied cohort.

|  | HC (N=20) | Chronic Pain (N=36) | Statistics<br>*Welch/t/χ<sup>2</sup>* | df | p-values |
| --- | --- | --- | --- | --- | --- |
| Age, in years, mean (SD) [range] | 50.48 (15.17) [24.2-80.86] | 52.17 (13.30) [23.77-74.81] | -0.431 | 54 | 0.668 |
| Sex, n (F/M) | 14/6 | 28/8 | 0.415 | 1 | 0.536 |
| PCL-C Total score (SD) [range] | 21.95 (5.10) [17-40] | 31.36 (9.63) [17-52] | **-4.780** | **53.94** | **<0.001** |
| PCL-C Re-experiencing score (SD) [range] | 6.15 (1.63) [5-11] | 7.81 (3.37) [5-17] | **-2.472** | **53.29** | **0.008** |
| PCL-C Avoidance score (SD) [range] | 8.80 (2.82) [7-19] | 13.06 (4.50) [7-28] | **-4.344** | **53.08** | **<0.001** |
| PCL-C Hyper-arousal score (SD) [range] | 7.00 (1.59) [5-10] | 10.50 (4.00) [5-19] | **-4.630** | **50.23** | **<0.001** |
| BDI total score (SD) [range]<sup>#</sup> | 3.61 (3.09) [0-10] | 10.40 (7.18) [0-29] | **-4.743** | **48.58** | **<0.001** |
| STAI-State, mean (SD) [range]<sup>§</sup> | 29.63 (6.80) [22-45] | 35.09 (8.69) [22-57] | **-2.351** | **50** | **0.011** |
| Pain condition<br>(TMD/TNP/BM/TRIG/TMD+TNP/SCI) | - | 14/8/1/6/1/6 | - | - | - |
| Pain duration, in years, mean (SD) [range]<sup>^</sup> | - | 10.03 (9.73) [1-36] | - | - | - |
| VAS pain diary, mean (SD) [range]<sup>%</sup> | - | 3.98 (2.21) [0.2-8.4] | - | - | - |
| VAS scan pain, mean (SD) [range]<sup>%</sup> | - | 3.56 (2.29) [0-9] | - | - | - |
| Scanning sites (NeuRA/SVH) | 14/6 | 29/6 | 1.233 | 1 | 0.319 |

HC: healthy controls; Pain: individuals with chronic pain; df: degrees of freedom; SD: standard deviation; F/M: females/males; BDI: Beck Depression Inventory; STAI: State-Trait Anxiety Inventory; TMD/TNP/BM/TRIG/TMD+TNP/SCI: temporomandibular disorder/trigeminal neuropathic pain/burning mouth/trigeminal neuralgia/temporomandibular disorder+trigeminal neuropathic pain/spinal cord injury; VAS: visual analogue scale; NeuRA:
Neuroscience Research Australia; SVH: St Vincent's Hospital Sydney
<sup>#</sup> Data missing for 2 HC and 2 chronic pain participants; <sup>§</sup> Data missing for 1 HC and 3 chronic pain participants; <sup>%</sup>: Data missing for 1 chronic pain participants; <sup>^</sup> missing data for 4 chronic pain participants
Significant group differences are in bold

### Magnetic Resonance Imaging

Imaging data were collected with two Phillips 3T Achieva TX scanner (Philips Healthcare, Best, The Netherlands) housed at Neuroscience Research Australia (Randwick, NSW, Australia) or at St Vincent’s Hospital (Darlinghurst, NSW, Australia), both equipped with 8-channel head coils. At both sites, a 3D T1-weighted structural scan covering the entire brain (acquisition parameters: repetition time (TR) = 5.6 ms, echo time (TE) = 2.5 ms, field of view (FOV) = 250 x 250 x 174 mm, 288 x 288 matrix, 200 sagittal slices, flip angle = 8°, voxel size = 0.9 x 0.9 x 0.9mm) and diffusion weighted imaging (DWI) were acquired (acquisition parameters: spin echo-planar imaging (EPI) acquisition sequence, 32 directions, b = 1000 s/mm2, TE/TR = 87/8879.79 ms, FOV = 224 x 137.5 x 224 mm, resolution = 2 mm isotropic).

A radiologist reviewed all scans before release to the investigators. Diffusion images were pre-processed following standard protocols from the Enhancing NeuroImaging Genetics through Meta-Analyses (ENIGMA) consortium (https://github.com/ENIGMA-git), using tools from the Functional MRI of the Brain Software Library (FSL; version 6.0.5.2) (26). Briefly, diffusion data were first denoised using the Marchenko-Pastur principal component analysis (MP-PCA) (27) and corrected for Gibbs ringing (28) using tools from the MRtrix3 software (respectively, *dwidenoise* and *mrdegibbs*) (29). Because only one phase encoding was collected, Synb0-DisCo (30) was used to create a synthetic b0 map from the T1- weighted image. Eddy currents were corrected using FSL *eddy*, and tensors were fitted with FSL *dtifit*. After each pre-processing step, the quality of images was assessed to identify potential errors in image processing using the ENIGMA guidelines. Finally, diffusion metrics such as fractional anisotropy (FA) and mean diffusivity (MD) were generated using FSL *tract-based spatial statistics* (TBSS) following the ENIGMA guidelines. Average FA and MD values were extracted for bilateral uncinate fasciculus, as well as the bilateral cingulate (CGC) and the hippocampal (CGH) portions of the cingulum.

### Harmonization

Before statistical analyses, extracted FA and MD values were harmonized using the python-based neuroHarmonize tools (https://github.com/rpomponio/neuroHarmonize) (31). Briefly, this approach uses empirical Bayes methods derived from the ‘ComBat’ R package (32) to adjust FA and MD values for the potential variations associated with the difference in scanning locations. To ensure neuroHarmonize does not remove the variance associated with age, sex, group, and PCL-C total score, these variables were added as covariates during harmonization.

### Statistical analyses

A series of multiple linear regressions were performed to determine the main effects of group (HC versus chronic pain), severity of PTSS (PCL-C total score) and their interaction (the product of group-by-mean-centred PCL-C total score), first on FA values of each a priori ROI (one model for each ROI), and second on MD values (one model for each ROI). Age and sex were added as covariates in all analyses.

For each index of diffusion separately (FA or MD), only models surviving Bonferroni correction to account for the number of ROIs (three: uncinate fasciculus, CGC, CGH) were considered (*p*=0.05/3=0.017). All analyses were performed in R (v4.3.1) (33) and RStudio (2023.6.2.561) (34).

In case of significant associations between the group-by-PTSS severity interaction and FA or MD, moderation analyses were performed using the ‘interactions’ R package (v1.1.5) (35). Group was used a priori as the moderator of the relationship between PTSS severity and indices of white matter microstructure integrity (FA or MD values). The Davidson–McKinnon correction (HC3) was used to account for heteroskedasticity (36) using the R package ‘sandwich’ (v3.2.2) (37, 38). Within each significant model, statistical significance was set at a threshold of *p*<0.05.

### Exploratory analyses

Symptom-specific effects were also explored using scores for the re-experiencing, avoidance and hyperarousal symptoms. Additional Bonferroni correction was applied to the original corrected threshold for significance to account for the number of symptoms studied for each ROIs (*p*=0.017/3=6x10^-3^).

## Results

### Participant characteristics

Demographic details are summarised in Table 1. Briefly, participants with chronic pain were not statistically different from the HC group in terms of age, sex and scanning site distributions. However, they reported more severe PTSS, depression, and anxiety than the HC group.

### Analyses for PTSS total score

Table 2 summarises the results of all statistical models tested (FA and MD). Analyses of FA indicated that models for the uncinate fasciculus and the CGC were significant, but only the model for the uncinate fasciculus survived Bonferroni correction (*p*<0.017). Within this model, and in the context of significant direct effects of trauma, the group-by-trauma interaction was significantly associated with variations of uncinate fasciculus FA. Moderation analysis indicated that increasing PTSS severity was significantly associated with decreased uncinate fasciculus FA in the HC group but not in the chronic pain group. Analyses of MD showed no statistically significant model.

**Figure 1.**
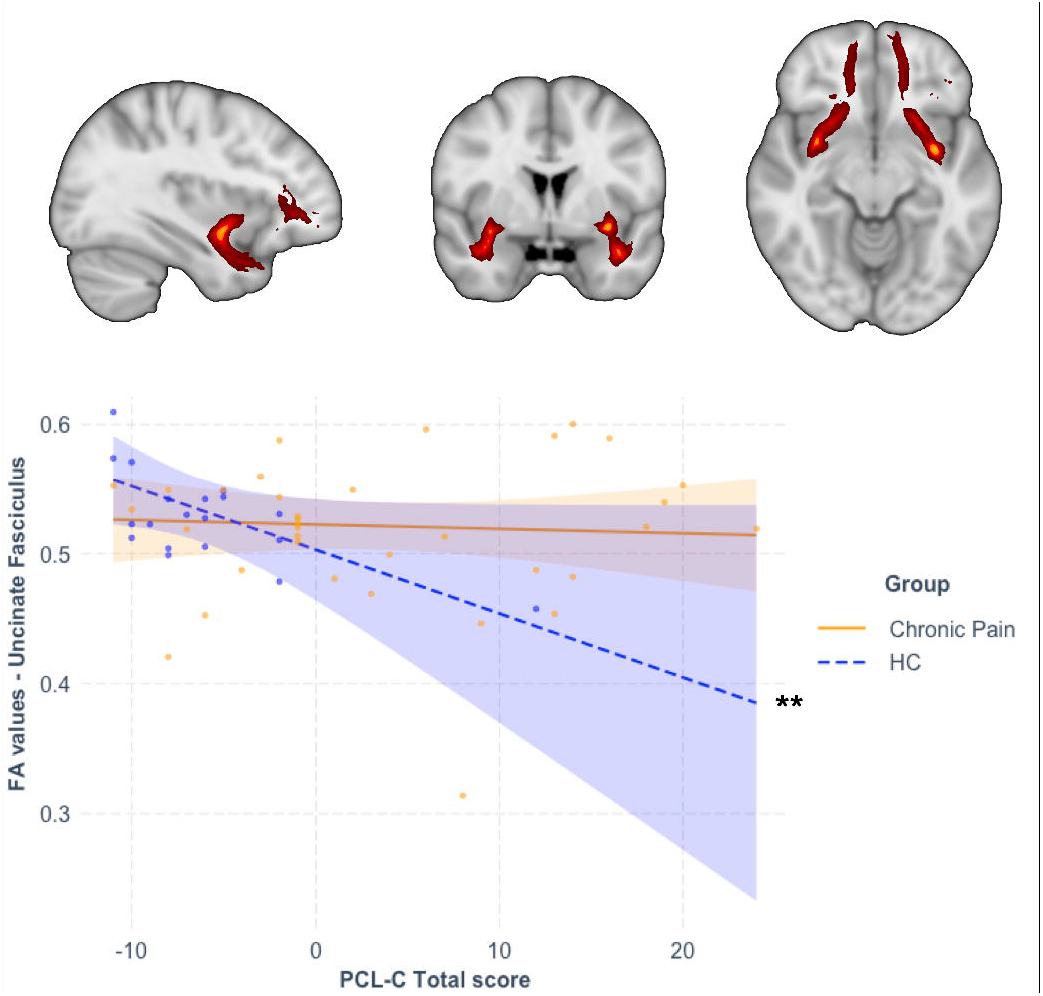
Moderation analysis following significant association between the group-by-PTSS interaction term and variations in fractional anisotropy values in the uncinate fasciculus. The interaction term was significantly associated with variations in fractional anisotropy (FA) in the uncinate fasciculus. Increasing severity of PTSS was significantly associated with decreased FA values in the uncinate fasciculus in the healthy control (HC group (blue dashed line), while this association was not significant in people with chronic pain (yellow plain line). ** *p*<0.01 Coloured band around each line represents 95% confidence intervals.

**Table 2.**
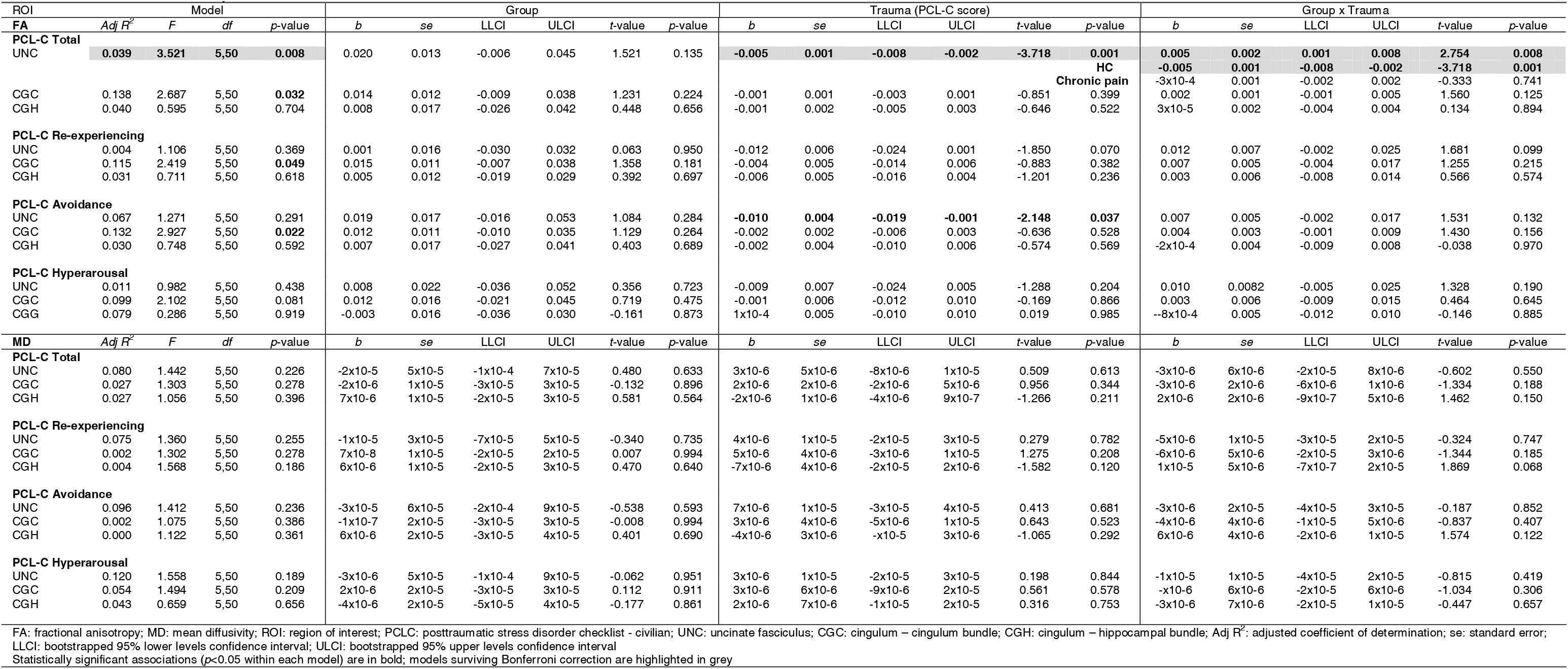
Results of the moderation analyses for all FA and MD ROIs.

### Analyses for specific PTSS

As shown in Table 2, the models investigating the moderating effect of group on the relationship between re-experiencing and avoidance symptoms and FA values in the CGC failed to survive Bonferroni correction (*p*<6x10^-3^). No other model for FA or MD values in relation to specific PTSS were significant.

## Discussion

The present study examined the differential relationship between comorbid PTSS and changes in white matter microstructural integrity in the cingulum and uncinate fasciculus, in people with and without chronic pain. Relative to the control group, the chronic pain group reported overall more severe PTSS. In addition, the group-by-PTSS severity interaction was significantly associated with changes in FA (but not MD) values in the uncinate fasciculus. Greater PTSS severity was associated with lower uncinate fasciculus integrity, only in the HC group. This was in the context of a direct association between PTSS severity and lower FA values in the uncinate fasciculus. There was no significant association between FA or MD values in any of the cingulum portions (cingulate or hippocampal) and group, PTSS severity of their interaction.

Partly consistent with our hypotheses, increased severity of PTSS was associated with decreased microstructural integrity of the uncinate fasciculus in the control group, but not in the chronic pain group. Reduced FA in the uncinate fasciculus is in line with previous reports in PTSD studies (39, 40). However, the lack of group difference or association between PTSS and FA values in the uncinate fasciculus in the chronic pain group was unexpected. Lower uncinate fasciculus FA associated with chronic pain is rather inconsistent, some groups reporting group differences (13) while others do not (41), indicating that specific pain types may have different effects on uncinate fasciculus integrity. One possible reason for the lack of association between uncinate fasciculus FA and PTSS severity in the chronic pain group, may be that the integrity of uncinate fasciculus, also reported in other conditions comorbid to chronic pain, may be mediated by these factors. For instance, depression has been found to impact the integrity of the uncinate fasciculus (42), and to affect the functional connectivity between the mPFC and the amygdala (43) two regions physically linked by the uncinate fasciculus, in chronic pain. Although depressive symptoms were measured, the relatively small sample size of this study prevented the investigation of cumulative effects of different psychopathological features on brain integrity. Future studies are warranted to disentangle these effects in larger samples.

Also surprising, was the lack of group differences and association with PTSS severity on cingulum integrity, either in its cingulate or hippocampal parts. In the context of the present chronic pain sample, it is possible that the lack of group difference in the cingulum may reflect the heterogeneity of the studied sample (9). Although we hypothesised that group differences would be evident across conditions, it is possible that the different types of pain (e.g., neuropathic versus musculoskeletal pain), or pain locations (e.g., localised versus widespread) may have different effects of brain white matter integrity. Studies of larger sample will help better identify the roles of these different factors. In addition, unlike for changes in grey matter integrity (7), the role of PTSS severity on white matter integrity in the cingulum may also depend on the timing of trauma exposure. A recent study showed that while decreased integrity of the uncinate fasciculus was associated with PTSS severity a month after trauma, decreased integrity of the cingulum was evident later, three months post-trauma (20). It is therefore possible that if people exposed to trauma do not develop clinical levels of PTSD, these changes in white matter may only be transient and normalise over time. This interpretation remains speculative, and longitudinal studies with both short (months) and long-term (years) outcomes are needed to confirm this.

This study has several limitations. First, the relatively small sample size may have limited our abilities to determine subtle effects and to investigate the roles of other psychopathological factors on white matter microstructural integrity. The effects of PTSS may be mediated by the severity of depressive symptoms in this population. Second, pharmacological treatments were not accounted for in the present analyses. This is partly due to the presence of the control group in all analyses and to the limited sample size. However, potential neurotrophic or neuroprotective effects of pharmacological treatments on brain morphology, function and neurochemistry (44) cannot be ruled out and must be acknowledged. Finally, although it can also be considered a strength, another potential limitation is the inclusion of people with different types (e.g., neuropathic, musculoskeletal) and location of pain (e.g., lower body, head) together. This approach is the most appropriate to identify changes in white matter microstructural integrity across pain conditions, but also increases the heterogeneity of the sample and may have contributed to confounding effects pertaining to specific pain types or locations.

In conclusion, partially consistent with our hypotheses, the severity of PTSS was associated with reduced white matter microstructural integrity (FA) in the uncinate fasciculus in people who do not experience chronic pain, but not in those experiencing chronic pain. There was no group difference or association with PTSS severity in the cingulum, or in mean diffusivity in any of the regions studied. These findings indicate that other factors, including but limiting to depressive symptoms, may be at play when investigating the effects of PTSS on brain integrity of people with chronic pain. Larger studies of more homogeneous samples are needed to better understand the interplay between trauma exposure, comorbid psychopathologies and brain integrity in people with chronic pain.

## Acknowledgments

The authors acknowledge the volunteers who participated in this study, and the assistance of previous students with data collection and entry and of medical personnel with participant recruitment.

## Data availability

All data produced in the present study are available upon reasonable request to the authors.

## Authors contributions

Y.Q. contributed conceptualisation, data curation, formal analysis, investigation, methodology, validation, visualisation, writing of the original draft, supervision, and review and editing. K.K. contributed conceptualisation, data curation, formal analysis, investigation, methodology, validation, visualisation, writing of the original draft, and review and editing. S.M.G. contributed conceptualisation, data curation, formal analysis, funding acquisition, investigation, methodology, project administration, resources, supervision, and review and editing.

